# Risk of Myocarditis and Pericarditis Following Coronavirus Disease 2019 Messenger RNA Vaccination—A Nationwide Study

**DOI:** 10.1101/2022.10.11.22280860

**Authors:** Wei-Ju Su, Yu-Lun Liu, Chia-Hsuin Chang, Yen-Ching Lin, Wei-I Huang, Li-Chiu Wu, Shu-Fong Chen, Yu-Sheng Lin, Yee-Lin Hsieh, Chiao-An Yang, Chiu-Hsiang Lin, Kim-Wei Arnold Chan, Ping-Ing Lee, Jen-Hsiang Chuang, Chin-Hui Yang

## Abstract

**Background:** An extended interval between the two primary doses may reduce the risk of myocarditis/pericarditis after COVID-19 mRNA vaccination. Taiwan has implemented a two-dose regimen with a 12-week interval for adolescents. Here we present nationwide data of mRNA COVID-19 vaccination-associated myocarditis and pericarditis in Taiwan.

**Methods:** Data on adverse events of myocarditis/pericarditis were from the Taiwan Vaccine Adverse Events Reporting System between March 22, 2021, and February 9, 2022. The rates according to sex, age, and vaccine type were calculated. We investigated the reporting rates among young individuals under different two-dose intervals and among those who received two doses of different vaccines.

**Results:** Among 204 cases who met the case definition of myocarditis/pericarditis, 75 cases occurred after the first dose and 129 after the second. The reporting rate of myocarditis/pericarditis after COVID-19 vaccination varied across sex and age groups and was highest after the second dose in males aged 12–17 years (126.79 cases per million vaccinees) for the BNT162b2 vaccine and in males aged 18–24 years (93.84 cases per million vaccinees) for the mRNA-1273 vaccine. The data did not suggest an association between longer between-dose interval and lower rate of myocarditis/pericarditis among males and females aged 18–24 or 25–29 years who received two doses of the BNT162b2 or mRNA-1273 vaccine. Rates of myocarditis/pericarditis in males and females aged 18–49 years after receiving ChAdOx1-S - mRNA-1273 vaccination was significantly higher than after ChAdOx1-S - ChAdOx1-S vaccination.

**Conclusions:** Myocarditis and pericarditis are rare following mRNA vaccination, with higher risk occurring in young males after the second dose.

## INTRODUCTION

After the implementation of mass COVID-19 vaccination programs in many countries, concern has been raised over an association between mRNA-based COVID-19 vaccines and acute myocarditis and pericarditis.^1^ In a meta-analysis of nine studies published in the English language, the overall risk of myopericarditis after mRNA vaccination was 22.6 cases (95% CI 12.2–42.0) per million doses, with the risk primarily driven by the higher incidence in young male recipients after the second dose of vaccine.^2^ However, the reported risks of myocarditis and pericarditis vary markedly.^3-5^ For example, after the second dose of BNT162b2 (Pfizer-BioNTech) vaccine, a Hong Kong study reported an incidence rate of myocarditis and pericarditis in males aged 12–17 years of 373.2 cases per million doses.^3^ An Israeli group reported an incidence rate of myocarditis following the second dose of the BNT162b2 vaccine in males aged 16–19 years as 150.7 cases per million doses.^4^ A U.S. study reported an incidence in males aged 16–17 years of 105.9 cases per million doses.^5^ The variations in risk may be related to differences in ethnicity, surveillance method, case definition, and observation period after vaccination across these studies. Some data also suggests that countries with an 8–12-week between-dose interval report a lower incidence of myocarditis after the second dose than countries with a 4-week between-dose interval.^6^

Since March 22, 2021, Taiwan has implemented a national immunization campaign. This campaign provided a first dose of ChAdOx1-S vaccine (Oxford/AstraZeneca) to 19,839,419 residents aged ≥ 18 years. Healthcare workers, public health workers, and non-healthcare frontline workers were the priority groups at the beginning of the COVID-19 vaccination program in Taiwan; vaccination was subsequently extended to those > 75 years of age, pregnant women, and vulnerable patients with underlying diseases.^7^ Later, the vaccines were sequentially offered to all residents aged ≥ 18 years in a schedule based primarily on age. Other COVID-19 vaccines became available later—mRNA-1273 vaccine (Moderna) on June 8, MVC-COV1901 vaccine (Medigen) on August 23, and BNT162b2 vaccine on September 22. The Advisory Committee on Immunization Practices (ACIP) in Taiwan recommends a minimum interval of 8 weeks between the first and second doses of the ChAdOx1-S and MVC-COV1901 vaccines, and a minimum of 4 weeks for the mRNA-1273 and BNT162b2 vaccines. From September 21, 2021, a school-based vaccination campaign for 1,218,167 teenagers aged 12–17 years was launched, in which the first dose of BNT162b2 vaccine was administered on campus. The administration of a second dose of BNT162b2 vaccine to teenagers was suspended on November 9, 2021, because of concern over an elevated risk of myocarditis. On November 29, 2021, ACIP revised the recommended two-dose interval for teenagers to 12 weeks, and vaccination was restarted on December 20, 2021.

In this study, we evaluated the rates of myocarditis and pericarditis following mRNA vaccination in Taiwan according to sex, age, vaccine type, and dose. The occurrence of myocarditis and pericarditis after homologous primary two-dose mRNA vaccination at different intervals and heterologous vaccination was also calculated.

## METHODS

### Data sources

#### COVID-19 vaccine safety surveillance in Taiwan

Based on vaccine safety surveillance of the mass immunization campaign during the influenza A (H1N1) pandemic in 2009, the Taiwanese government developed a strategy for timely assessment of vaccine safety.^8^ Adverse events following immunization (AEFI) surveillance is jointly operated by the Taiwan Center for Disease Control (TCDC) and the Taiwan Food and Drug Administration (TFDA). Previously, the information was collected on paper and entered into the Taiwan National Adverse Drug Reaction (ADR) Reporting system, which is operated by the Taiwan Drug Relief Foundation (TDRF). To monitor COVID-19 vaccine safety more efficiently, in addition to the established surveillance system, in 2021 the TCDC initiated the web-based Vaccine Adverse Report System (VAERS). VAERS accepts adverse events reported mainly by health care providers, contracted health facilities, and local health authorities. For COVID-19 vaccine safety surveillance, all AEFI reports received by VAERS following COVID-19 vaccination were screened, and reported adverse events were assigned a Medical Dictionary for Regulatory Activities (MedDRA) code.

Suspected myocarditis/pericarditis reports were identified by searching using the MedDRA preferred term (PT) of myocarditis or pericarditis. These reports, in addition to the medical records (when available), were reviewed by TDRF staff to determine the level of diagnostic certainty using the Brighton Collaboration Case Definition for myocarditis/pericarditis.^9^ Reports that met level 1–3 of the case definition were considered potential cases. Reports with insufficient evidence to meet level 1–3 of the case definition were re-examined when additional information was received.

#### National Immunization Information System

We obtained the numbers of persons receiving COVID-19 vaccinations between March 22, 2021, and February 9, 2022, from the web-based National Immunization Information System (NIIS), established by the TCDC in 2003. Electronic records for all publicly funded vaccines were compiled for all citizens of Taiwan born after 1980. According to a TCDC survey of the NIIS database, the accuracy of publicly funded immunization electronic records was > 90%. NIIS accepts daily electronic reports on COVID-19 vaccines administered at all contracted healthcare facilities and vaccination sites. The datasets comprise information on date of vaccine administration, target group, vaccine manufacturer, and vaccine dose. NIIS was used for timely monitoring of vaccine coverage among priority groups at the national and county levels, and to provide denominator data for calculating rates of adverse events among vaccine recipients.

#### Outcome definitions

We evaluated the occurrence of myocarditis and pericarditis after receiving a first and second dose of mRNA COVID-19 vaccine. We obtained data from myocarditis and pericarditis cases reported to VAERS between March 22, 2021, and February 9, 2022, and confirmed by the ADR center according to the Brighton criteria for myocarditis/pericarditis level 1–3 without restriction on time of onset after vaccination. We classified patients with myocarditis/pericarditis by sex, age (12–17, 18–24, 25–29, 30–39, 40–49, 50–59, 60–69, 70–79, and ≥ 80 years), vaccine type (BNT162b2 or mRNA-1273), and dose (first or second).

#### Statistical analysis

In the descriptive analysis, we used frequencies, percentages, means, and medians to characterize the distribution of demographic and clinical variables among cases. For each sex and age group, we calculated the crude reporting rate by dividing the number of cases of myocarditis/pericarditis by the number of vaccine recipients and computed the 95% confidence intervals by exact Poisson distribution. We investigated rates of myocarditis/pericarditis among young individuals after two doses of the same mRNA vaccine at an interval of ≤ 28, 29–55, 56–83, or ≥ 84 days and among those who received two doses of different vaccines.

#### Ethics

Because this study was a component of the public health response function of the Central Epidemic Command Center for surveillance purposes, it received a waiver for review by an institutional review board. Information was collected according to the regulations of the Central Epidemic Command Center and in accordance with Article 17 of the Communicable Disease Control Act. The data were de-identified before analysis.

## RESULTS

As of February 9, 2022, a total of 4,015,618 persons aged ≥ 18 years had received a first dose and 3,566,249 persons had received first and second doses of mRNA-1273 vaccine. For individuals aged ≥ 12 years, 6,310,060 had received a first dose and 5,497,108 had received first and second doses of BNT162b2 vaccine. Table 1 lists mRNA vaccination rates according to sex, age, vaccine type, and vaccine dose.

**Table 1.** Number of mRNA COVID-19 vaccine recipients, number of myocarditis and pericarditis cases after vaccination, and risk of myocarditis and pericarditis in Taiwan,, March 22, 2021-Febuary 9, 2022

|  | BNT162b2 vaccine |  |  |  |  |  | mRNA-1273 vaccine |  |  |  |  |  |
| --- | --- | --- | --- | --- | --- | --- | --- | --- | --- | --- | --- | --- |
|  | First dose |  |  | Second dose |  |  | First dose |  |  | Second dose |  |  |
| Age | No. of cases | No. of people received vaccination | Risk per million vaccinees | No. of cases | No. of people received vaccination | Risk per million vaccinees | No. of cases | No. of people received vaccination | Risk per million vaccinees | No. of cases | No. of people received vaccination | Risk per million vaccinees |
| Male |  |  |  |  |  |  |  |  |  |  |  |  |
| 12-17 | 25 | 579,277 | 43.16<br>(27.93~63.71) | 65 | 512,673 | 126.79<br>(97.85~161.60) | - | - | - | - | - | - |
| 18-24 | 4 | 485,820 | 8.23<br>(2.24~21.08) | 12 | 401,881 | 29.86<br>(15.43~52.16) | 1 | 74,979 | 13.34<br>(0.34~74.31) | 5 | 53,282 | 93.84<br>(30.47~218.99) |
| 25-29 | 1 | 319,788 | 3.13<br>(0.08~17.42) | 5 | 261,381 | 19.13<br>(6.21~44.64) | 2 | 87,876 | 22.76<br>(2.76~82.21) | 5 | 63,657 | 78.55<br>(25.50~183.30) |
| 30-39 | 3 | 592,463 | 5.06<br>(1.04~14.80) | 2 | 498,535 | 4.01<br>(0.49~14.49) | 4 | 172,390 | 23.20<br>(6.32~59.41) | 3 | 129,063 | 23.24<br>(4.79~67.93) |
| 40-49 | 3 | 558,469 | 5.37<br>(1.11~15.70) | 1 | 480,477 | 2.08<br>(0.05~11.60) | 1 | 169,271 | 5.91<br>(0.15~32.92) | 1 | 129,648 | 7.71<br>(0.20~42.98) |
| 50-59 | 0 | 390,300 | 0 | 0 | 341,441 | 0 | 1 | 256,434 | 3.90 | 1 | 219,993 | 4.55 |
|  |  |  |  |  |  |  |  |  | (0.10~21.73) |  |  | (0.12~25.33) |
| 60-69 | 0 | 171,997 | 0 | 0 | 147,953 | 0 | 2 | 636,202 | 3.14<br>(0.38~11.36) | 0 | 596,343 | 0 |
| 70-79 | 0 | 18,173 | 0 | 0 | 12,962 | 0 | 0 | 438,111 | 0 | 1 | 417,076 | 2.40<br>(0.06~13.36) |
| ≥ 80 | 0 | 6,245 | 0 | 0 | 4,094 | 0 | 0 | 62,620 | 0 | 0 | 54,811 | 0 |
| ≥ 18 | 11 | 2,543,255 | 4.33<br>(2.16~7.74) | 20 | 2,148,724 | 9.31<br>(5.69~14.38) | 11 | 1,897,883 | 5.80<br>(2.89~10.37) | 16 | 1,663,873 | 9.62<br>(5.50~15.62) |
| Female |  |  |  |  |  |  |  |  |  |  |  |  |
| 12-17 | 9 | 536,822 | 16.77<br>(7.67~31.83) | 8 | 486,145 | 16.46<br>(7.10~32.42) | - | - | - | - | - | - |
| 18-24 | 3 | 452,567 | 6.63<br>(1.37~19.37) | 4 | 406,764 | 9.83<br>(2.68~25.18) | 0 | 67,164 | 0 | 0 | 54,057 | 0 |
| 25-29 | 0 | 285,141 | 0 | 0 | 250,569 | 0 | 0 | 93,147 | 0 | 0 | 75,930 | 0 |
| 30-39 | 4 | 587,668 | 6.81<br>(1.85~17.43) | 3 | 518,541 | 5.79<br>(1.19~16.91) | 0 | 209,283 | 0 | 3 | 173,502 | 17.29<br>(3.57~50.53) |
| 40-49 | 2 | 631,025 | 3.17<br>(0.38~11.45) | 0 | 560,492 | 0 | 0 | 164,660 | 0 | 0 | 132,815 | 0 |
| 50-59 | 2 | 448,722 | 4.46<br>(0.54~16.10) | 1 | 403,282 | 2.48<br>(0.06~13.82) | 1 | 255,416 | 3.92<br>(0.10~21.81) | 1 | 224,045 | 4.46<br>(0.11~24.87) |
| 60-69 | 3 | 209,275 | 14.34<br>(2.96~41.89) | 0 | 184,266 | 0 | 2 | 720,132 | 2.78<br>(0.34~10.03) | 5 | 675,477 | 7.40<br>(2.40~17.27) |
| 70-79 | 0 | 26,085 | 0 | 0 | 18,850 | 0 | 2 | 521,504 | 3.84<br>(0.46~13.85) | 3 | 492,263 | 6.09<br>(1.26~17.81) |
| ≥ 80 | 0 | 10,223 | 0 | 0 | 6,802 | 0 | 0 | 86,429 | 0 | 0 | 74,287 | 0 |
| ≥ 18 | 14 | 2,650,706 | 5.28<br>(2.89~8.86) | 8 | 2,349,566 | 3.40<br>(1.47~6.71) | 5 | 2,117,735 | 2.36<br>(0.77~5.51) | 12 | 1,902,376 | 6.31<br>(3.26~11.02) |

### Characteristics of patients with myocarditis/pericarditis following mRNA vaccination

Between March 22, 2021, and February 9, 2022, VARES received a total of 5,957 and 2,408 reports of adverse events after the first and second doses of COVID-19 vaccine. Among the 238 reports of adverse events of special interest (AESI) that met Brighton criteria levels 1–3 for myocarditis or pericarditis, 189 (79.4%) were myocarditis (including myopericarditis) and 49 (20.6%) were pericarditis alone. Of these patients, 203 (85.3%) required hospitalization and 101 (age range 12–85 years) were admitted to the ICU. Of the 101 ICU patients, 9 were placed on extracorporeal membrane oxygenation and four died (age 39–76 years). Myocarditis/pericarditis occurred in 90 patients after the first dose of vaccine and in 148 after the second dose (138 underwent homologous and 10, heterologous vaccination). Of the 228 patients who developed myocarditis/pericarditis after the first or second dose of the same vaccine, 161 (70.6%) were male and their age was 12–85 years (mean 26.9 years; median 19 years). Symptoms manifested 0–71 days after vaccination (median 4 days), with 156 (68.4%) occurring 0–7 days after vaccination, 47 (20.6%) 8–28 days after vaccination, and 25 (11.0%) 29 days after vaccination. Among the 138 cases of myocarditis/pericarditis after receiving the second dose of the same vaccine, the between-dose interval was 29–116 days. Among them, 1 patient had a two-dose interval of < 30 days, 25 patients of ≤ 45 days, 50 patients of 46–90 days, and 63 patients an interval of > 90 days.

### Myocarditis/pericarditis rates according to sex, age, vaccine type, and vaccine dose

Table 1 lists the crude reporting rates of myocarditis/pericarditis according to sex, age, vaccine type, and vaccine dose. For the BNT162b2 vaccine, among individuals aged ≥ 18 years, the rate of myocarditis/pericarditis after the first dose was 4.33 and 5.28 cases per million vaccinees in males and females, respectively. However, the rates after the second dose in males and females were 9.31 and 3.40 cases per million vaccinees, respectively. Among all sex and age groups, the highest reporting rate of myocarditis/pericarditis was in males aged 12–17 years after the first dose (43.16 cases per million vaccinees) and after the second dose (126.79 cases per million vaccinees), followed by men aged 18–24 years after the second dose (29.86 cases per million vaccinees), and in males aged 25–29 years after the second dose (19.13 cases per million vaccinees). For females, the highest reporting rate was in those aged 12–17 years (16.77 and 16.46 cases per million vaccinees after the first and second doses, respectively). The rate of myocarditis/pericarditis in females aged 12–29 years was unchanged after the second dose of BNT162b2 vaccine.

For the mRNA-1273 vaccine, the crude reporting rate of myocarditis/pericarditis in males and females aged ≥ 18 years was 5.80 and 9.62, and 2.36 and 6.31 cases per million vaccinees after the first and second doses, respectively. The highest rate of myocarditis/pericarditis was in males aged 18–24 years (13.34 and 93.84 cases per million vaccinees after the first and second doses, respectively), followed by males aged 25–29 years (22.76 and 78.55 cases per million vaccinees after the first and second doses, respectively) and males aged 30–39 years (23.20 and 23.24 cases per million vaccinees after the first and second doses, respectively). The occurrence of myocarditis/pericarditis in young males was higher after receiving the mRNA-1273 vaccine than the BNT162b2 vaccine. A small number of women aged 18–29 years received the mRNA-1273 vaccine; no case of myocarditis/pericarditis was reported in this group. However, the rates of myocarditis/pericarditis were higher in women > 30 years of age after a second dose of the mRNA-1273 vaccine than after a second dose of the BNT162b2 vaccine. Rates of myocarditis/pericarditis following ChAdOx1-S and MVC-COV1901 vaccination are listed in Supplementary Table.

### Risks after receiving two doses of the same vaccine at different between-dose intervals and after two doses of different vaccines

Table 2 lists the rates of myocarditis/pericarditis after two doses of the same mRNA vaccine among individuals aged 12–29 years stratified by between-dose interval (≤ 28, 29–55, 56–83, and ≥ 84 days). The data did not suggest an association between longer between-dose interval and lower rate of myocarditis/pericarditis among males and females aged 18–24 or 25–29 years who received two doses of the BNT162b2 or mRNA-1273 vaccine. However, these data should be interpreted cautiously because of the small number of vaccine recipients.

**Table 2.**
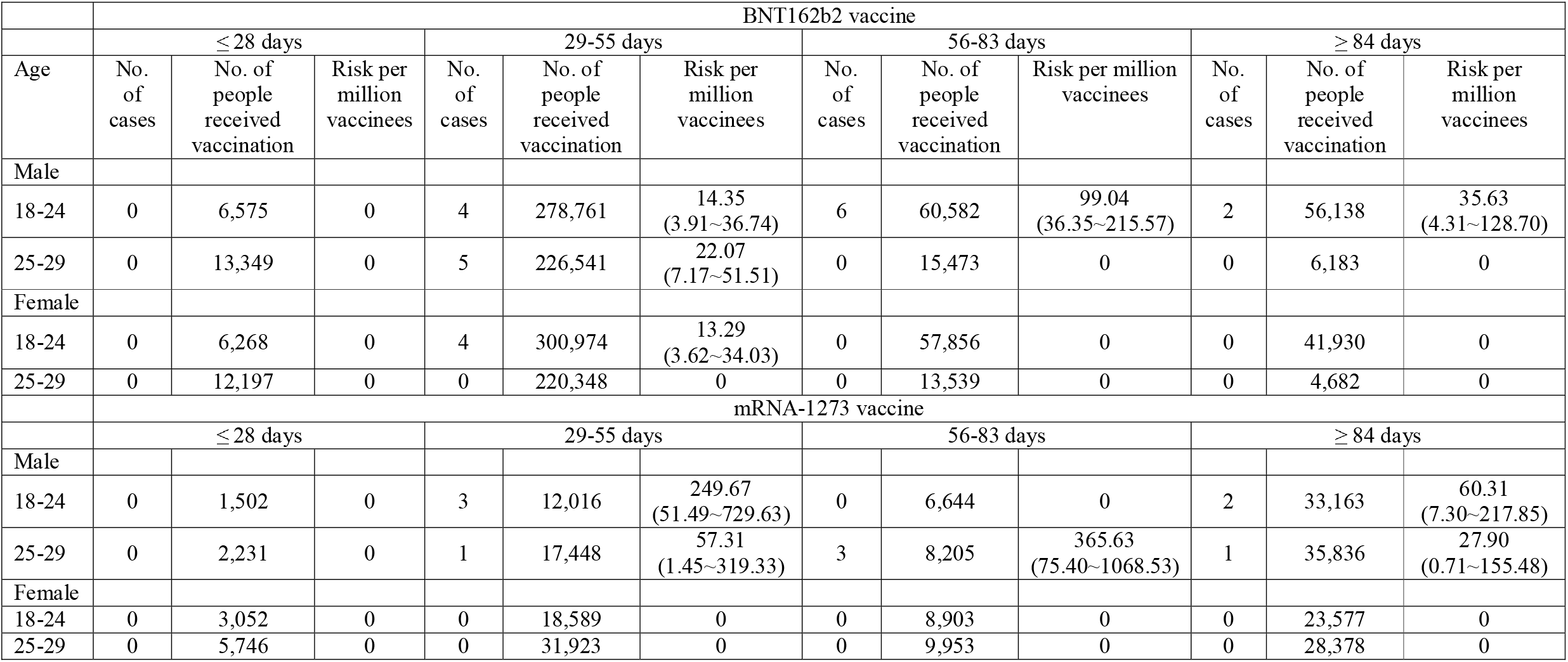
Number of two doses of BNT162b2 and mRNA-1273 COVID-19 vaccine recipients, number of myocarditis and pericarditis cases after vaccination, and risk of myocarditis/pericarditis stratified on the interval between the first and second dose of vaccination among young population in Taiwan, March 22, 2021-Febuary 9, 2022

As of February 9, 2022, the ADR center affirmed that 10 patients developed myocarditis/pericarditis after receiving two doses of different vaccines (8 myocarditis and 2 pericarditis). Among them, five were males, aged 18–38 years. Nine patients received ChAdOx1-S as the first dose and mRNA-1273 as the second dose; one received ChAdOx1-S as the first dose and BNT162b2 as the second dose. The rates of myocarditis/pericarditis among patients who received two doses of different vaccines are listed in Table 3. The occurrence of myocarditis/pericarditis after receiving ChAdOx1-S - mRNA-1273 vaccination was 81.3 cases per million vaccinees in males aged 18-24 years, approximately 40 cases per million vaccinees in males aged 25-39 years, and about 30 cases per million vaccinees in females aged 18-49 years, significantly higher than after ChAdOx1-S - ChAdOx1-S vaccination.

**Table 3.**
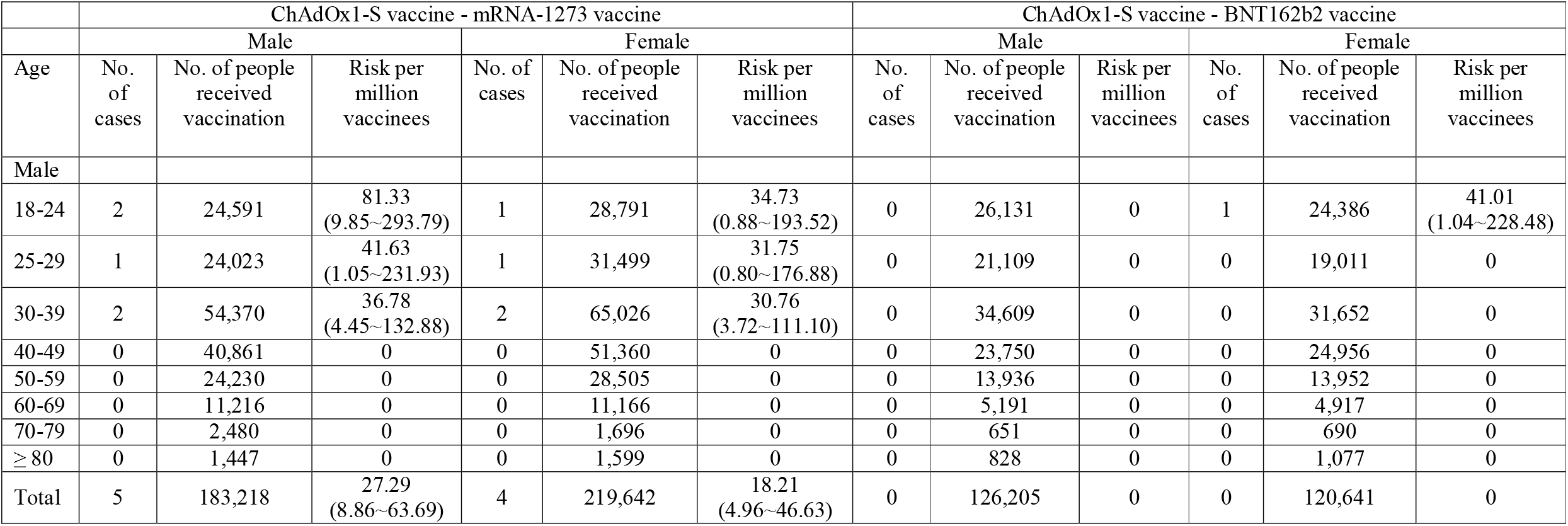
Number of heterologous ChAdOx1-S vaccine and mRNA COVID-19 vaccine recipients, number of myocarditis and pericarditis cases after vaccination, and risk of myocarditis/pericarditis in Taiwan, March 22, 2021-Febuary 9, 2022

## DISCUSSION

We found substantial variations in the reporting rates of myocarditis and pericarditis after mRNA vaccination according to sex and age, and vaccine dose. The absolute risks of myocarditis/pericarditis following mRNA vaccination were higher in males than in females and after the second compared to after the first dose, and decreased with age. For most age groups and both sexes, the occurrence of myocarditis and of pericarditis following mRNA vaccination were low, with the highest absolute risk occurring in young males after the second dose of mRNA vaccine. These findings are consistent with prior reports.

The reporting rates of myocarditis following mRNA vaccination varied widely among population-based studies. Some data suggests that countries with an 8–12-week between-dose interval report a lower incidence of myocarditis after the second dose than countries with a 4-week between-dose interval.^6^ Supporting data comes from the preliminary results of a Canadian study, which indicate that among males aged 18–24 years receiving two doses of the same mRNA vaccine, the rate of myocarditis was lower in recipients with a > 56-day compared to a < 30-day between-dose interval.^10^ However, under the Taiwanese policy of a 12-week between-dose interval, the rate of myocarditis in males aged 12–17 years after the second dose of BNT162b2 vaccine appeared not to be lower than in the U.S.^5^ and Israel^4^ with a 3-week between-dose interval. Based on passive surveillance reporting in Taiwan, the data did not suggest an association between longer between-dose interval of BNT1626b vaccine and lower rate of myocarditis in males aged 18–29 years. More high-quality data are required to conclude whether extending the between-dose interval from 4 to 8 or 12 weeks would reduce the incidence of myocarditis.

Myocarditis/pericarditis after mRNA vaccination is rare in Asian populations, although the rates vary by gender and age group. For the highest-risk group (males aged 12–17 years), the occurrence of myocarditis/pericarditis after the second dose of mRNA vaccine is about 1 in 10,000. Among young patients who developed myocarditis following mRNA vaccination, only one had severe disease and none died. There is much evidence that the overall benefits of COVID-19 vaccination far exceed the potential harm caused by adverse reactions. Although the clinical symptoms of severe acute respiratory syndrome coronavirus-2 (SARS-CoV-2) infection tend to decrease in severity with the emergence of new mutants, vaccination of the entire population is critical for controlling the pandemic. The safety information of a new vaccine is crucial for its acceptance by the population.

Cases of myocarditis caused by mRNA vaccines are likely to be over-reported on social media in the initial stages of a mass vaccination campaign, and no country has sufficient data to assess accurately the risks and benefits of the vaccines. An automated active surveillance system that pools data from multiple electronic healthcare databases from several countries could monitor vaccine-related adverse events in real time. A common protocol with unified monitoring methods, case definitions, analysis plans, reporting, and data sharing, would yield accurate information to support vaccine policymaking and risk management and increase public confidence in vaccine safety.

Our results are based on the population-based data of almost 20 million vaccinated individuals in Taiwan. We did not restrict the observation time after vaccination to collect all mRNA vaccine-related myocarditis and pericarditis cases. The level of diagnostic certainties of myocarditis/pericarditis cases was confirmed by the clinical pharmacists at the National Adverse Drug Reaction Center. We focus on absolute risk as it provides the estimate of disease burden related to vaccine-associated myocarditis. We modeled risk separately after the first and second doses of vaccine according to sex, age, and vaccine type, and calculated the risks after two doses of the same vaccine at different between-dose intervals and after two doses of different vaccines. There was no significant COVID-19 outbreak in Taiwan during the study period, reducing the likelihood of misclassification of the cause of myocarditis as the vaccine or SARS-CoV-2 infection.

This study had several limitations. First, because VARES is a passive surveillance system, we could not exclude the possibility that healthcare providers did not report some potential cases to VARES. However, given the many reports of myocarditis and pericarditis in foreign literature, health care providers in Taiwan are well aware of this outcome and extent of under-reporting would be small. Second, because the number of young people receiving mRNA-1273 vaccination was small, we could not estimate precisely the incidence of mRNA-1273-associated myocarditis. Third, because the numbers of individuals receiving BNT162b2 and mRNA-1273 vaccination varied markedly, probably due to differential availability, we did not directly compare their risks of myocarditis/pericarditis.

In conclusion, myocarditis and pericarditis are rare following mRNA vaccination, with higher occurrence occurring in young males after the second dose of mRNA vaccine. The benefits of COVID-19 vaccination far outweigh the possible harm for either the individual or society.

## Supporting information

Myocarditis/pericarditis after COVID-19 vaccination in Taiwan - Supplementary files

## Data Availability

All data produced in the present work are contained in the manuscript.

## Author contributions

Drs. Su, Liu, and Yang had full access to all of the data in the study and take responsibility for the integrity of the data and the accuracy of the data analysis.

*Concept and design:* Su, Liu, Chang, and Yang

*Acquisition, analysis, or interpretation of data:* Su, Liu, Wu, Lin, Chang and Chan

*Drafting of the manuscript:* Su, Lin, and Chang

*Critical revision of the manuscript for important intellectual content:* Liu, Huang, Chen, Lin, Hsieh, Yang, Lin, Chan, Lee, Chuang, and Yang,

*Administrative, technical, or material support:* Su, Liu and Yang

*Supervision:* Chan, Lee, Chuang, and Yang

All the authors have read, contributed to, and approved the final manuscript and agreed to transfer the copyright ownership in the event of acceptance.

## Conflict of Interest Disclosure

None reported.

## Grant support

The study is not supported by any funding resources.

## Notes

### Competing Interest Statement

The authors have declared no competing interest.

### Funding Statement

This study did not receive any funding

### Author Declarations

The Institutional Review Board of Centers for Disease Control, Ministry of Health and Welfare, Taiwan, waived ethical approval for this work.

## REFERENCES

1. Bozkurt B, Kamat I, Hotez PJ. Myocarditis With COVID-19 mRNA Vaccines. Circulation 2021;144:471–84.

2. Ling RR, Ramanathan K, Tan FL, Tai BC, Somani J, Fisher D, et al. Myopericarditis following COVID-19 vaccination and non-COVID-19 vaccination: a systematic review and meta-analysis. Lancet Respir Med 2022;10:679–88.

3. Chua GT, Kwan MYW, Chui CSL, Smith RD, Cheung EC, Tian T, et al. Epidemiology of Acute Myocarditis/Pericarditis in Hong Kong Adolescents Following Comirnaty Vaccination. Clin Infect Dis 2021 Nov 28.

4. Mevorach D, Anis E, Cedar N, Bromberg M, Haas EJ, Nadir E, et al. Myocarditis after BNT162b2 mRNA Vaccine against Covid-19 in Israel. N Engl J Med 2021;385:2140–9.

5. Oster ME, Shay DK, Su JR, Gee J, Creech CB, Broder KR, et al. Myocarditis Cases Reported After mRNA-Based COVID-19 Vaccination in the US From December 2020 to August 2021. JAMA 2022;327:331–40.

6. Moulia D. Myocarditis and COVID-19 vaccine intervals : international data and policies. In: United States. Advisory Committee on Immunization Practices C-VWG, editor.; 2022; Atanta, GA; 2022.

7. Taiwan Centers for Disease Control. COVID-19 vaccine priority groups. Accessed May 5, 2022. Available from: https://www.cdc.gov.tw/Category/Page/9mcqWyq51P_aYADuh3rTBA.

8. Huang WT, Chen WW, Yang HW, Chen WC, Chao YN, Huang YW, et al. Design of a robust infrastructure to monitor the safety of the pandemic A(H1N1) 2009 vaccination program in Taiwan. Vaccine 2010;28:7161–6.

9. Brighton Collaboration. Myocarditis/Pericarditis Case Definition. Accessed May 5, 2022. Available from: https://brightoncollaboration.us/myocarditis-case-definition-update/.

10. Buchan SA, Seo CY, Johnson C, Alley S, Kwong JC, Nasreen S, et al. Epidemiology of Myocarditis and Pericarditis Following mRNA Vaccination by Vaccine Product, Schedule, and Interdose Interval Among Adolescents and Adults in Ontario, Canada. JAMA Network Open 2022;5:e2218505–e.

