## Supplementary material for "Risk of Myocarditis and Pericarditis Following Coronavirus Disease 2019 Messenger RNA Vaccination—A Nationwide Study": Myocarditis/pericarditis after COVID-19 vaccination in Taiwan - Supplementary files

**Supplementary Table.** Number of ChAdOx1-Sand MVC-COV1901COVID-19 vaccine recipients, number of myocarditis and pericarditis cases after vaccination, and risk of myocarditis and pericarditis in Taiwan, 2021/3/22-2022/2/9

| | ChAdOx1-S vaccine | | | | | | MVC-COV1901 vaccine | | | | | |
|---|---|---|---|---|---|---|---|---|---|---|---|---|
| | First dose | | | Second dose | | | First dose | | | Second dose | | |
| Age | No. of cases | No. of people received vaccination | Risk per million vaccinees | No. of cases | No. of people received vaccination | Risk per million vaccinees | No. of cases | No. of people received vaccination | Risk per million vaccinees | No. of cases | No. of people received vaccination | Risk per million vaccinees |
| Male | | | | | | | | | | | | |
| 12-17 | - | - | - | - | - | - | - | - | - | - | - | - |
| 18-24 | 1 | 346,854 | 2.88 (0.07~16.06) | 0 | 269,163 | 0 | 0 | 37,185 | 0 | 0 | 32,113 | 0 |
| 25-29 | 2 | 362,058 | 5.52 (0.67~19.95) | 2 | 293,459 | 6.82 (0.83~24.62) | 0 | 48,338 | 0 | 0 | 42,351 | 0 |
| 30-39 | 2 | 750,993 | 2.66 (0.32~9.62) | 1 | 625,444 | 1.60 (0.04~8.91) | 0 | 113,410 | 0 | 0 | 100,745 | 0 |
| 40-49 | 1 | 811,278 | 1.23 (0.03~6.87) | 0 | 717,578 | 0 | 0 | 97,253 | 0 | 0 | 83,373 | 0 |
| 50-59 | 1 | 762,624 | 1.31 (0.03~7.31) | 0 | 701,679 | 0 | 0 | 74,825 | 0 | 0 | 64,184 | 0 |

| | | | | | | | | | | | | |
|---|---|---|---|---|---|---|---|---|---|---|---|
| 60-69 | 1 | 412,871 | 2.42 (0.06~13.49) | 1 | 383,393 | 2.61 (0.07~14.53) | 0 | 51,117 | 0 | 0 | 43,611 | 0 |
| 70-79 | 0 | 143,864 | 0 | 0 | 135,016 | 0 | 0 | 8,722 | 0 | 0 | 6,722 | 0 |
| ≥ 80 | 1 | 202,259 | 4.94 (0.13~27.55) | 0 | 180,189 | 0 | 0 | 2,742 | 0 | 0 | 1,859 | 0 |
| Total | 9 | 3,792,801 | 2.37 (1.09~4.50) | 4 | 3,305,921 | 1.21 (0.33~3.10) | 0 | 433,592 | 0 | 0 | 374,958 | 0 |
| Female | | | | | | | | | | | |
| 12-17 | - | - | | - | - | - | - | - | - | - | - | - |
| 18-24 | 1 | 353,047 | 2.83 (0.07~15.78) | 2 | 284,495 | 7.03 (0.85~25.39) | 0 | 36,751 | 0 | 0 | 33,409 | 0 |
| 25-29 | 1 | 382,990 | 2.61 (0.07~14.55) | 1 | 317,089 | 3.15 (0.08~17.57) | 0 | 45,079 | 0 | 0 | 41,114 | 0 |
| 30-39 | 1 | 812,766 | 1.23 (0.03~6.86) | 0 | 686,707 | 0 | 0 | 103,184 | 0 | 0 | 93,986 | 0 |
| 40-49 | 2 | 977,281 | 2.05 (0.25~7.39) | 1 | 875,563 | 1.14 (0.03~6.36) | 0 | 87,000 | 0 | 1 | 76,874 | 13.01 (0.33~72.48) |
| 50-59 | 0 | 866,933 | 0 | 0 | 806,071 | 0 | 0 | 64,555 | 0 | 0 | 56,300 | 0 |
| 60-69 | 0 | 442,727 | 0 | 0 | 415,595 | 0 | 1 | 43,110 | 23.20 (0.59~129.24) | 0 | 36,730 | 0 |
| 70-79 | 0 | 153,638 | 0 | 0 | 145,402 | 0 | 0 | 8,075 | 0 | 0 | 5,898 | 0 |
| ≥ 80 | 0 | 266,919 | 0 | 0 | 236,912 | 0 | 0 | 3,799 | 0 | 0 | 2,507 | 0 |

| | | | | | | | | | | | | |
|---|---|---|---|---|---|---|---|---|---|---|---|---|
| Total | 5 | 4,256,301 | 1.17<br>(0.38~2.74) | 4 | 3,767,834 | 1.06<br>(0.29~2.72) | 1 | 391,553 | 2.55<br>(0.06~14.23) | 1 | 346,818 | 2.88<br>(0.07~16.07) |
